# Federated Learning for Multi-Disease Ophthalmic Diagnostics using OCTA

**DOI:** 10.1101/2025.04.25.25326431

**Authors:** Ahammed Sakir Nabil, Sina Gholami, Theodore Leng, Jennifer I Lim, Minhaj Nur Alam

## Abstract

Federated learning enables collaborative model training across multiple institutions while preserving patient data privacy. This study evaluates five different aggregation strategies (FedAvg, FedAdagrad, FedYogi, FedProx, and FedMRI) for federated learning in the context of multi-disease retinal disease classification using optical coherence tomography angiography (OCTA). We tested these approaches on a diverse dataset combining public OCTA-500 and private data provided by the University of Illinois Chicago (UIC) across seven distinct retinal pathologies, comparing performance against centralized and standalone models in three experimental scenarios of varying class complexity. Our results demonstrate that federated approaches can match or even exceed centralized training performance, with FedMRI achieving 60.87% accuracy in the comprehensive seven-class scenario and all three primary federated methods (FedAvg, FedProx, FedMRI) outperforming centralized training in simplified class scenarios (72.09% vs 69.77%). We observed that different aggregation strategies excel in different performance metrics—FedMRI consistently demonstrated superior ROC-AUC performance while FedAvg showed stronger F1-scores, suggesting better class balance management. These findings provide practical insights for implementing privacy-preserving collaborative AI systems in OCTA-based ophthalmic diagnostics.

## I. INTRODUCTION

Medical imaging has revolutionized healthcare by enabling non-invasive visualization of internal structures. It accounts for almost 90% of all healthcare data [1] and thus, plays a pivotal role in diagnosis, treatment planning, and disease monitoring [2]. The continuous evolution of imaging technologies, from X-rays to advanced molecular imaging [3], has dramatically improved our ability to detect and characterize pathologies at earlier stages, leading to better patient outcomes. In ophthalmology, this evolution has been particularly significant, as the intricate structure of the eye demands high-resolution imaging for accurate diagnosis and treatment planning [4].

Vision impairment and blindness affect approximately 2.2 billion people globally, with retinal diseases such as Diabetic Retinopathy (DR) being primary contributors [5]. The key to preventing irreversible vision loss lies in early detection and accurate diagnosis, particularly as the global burden of these diseases is expected to increase with aging populations and rising diabetes prevalence [6]. However, the subtle variations in retinal pathologies make diagnosis particularly challenging, even for experienced clinicians [7].

Traditional ophthalmological imaging modalities such as fundus photography and fluorescein angiography (FA) have long served as standard tools for retinal examination. While fundus photography provides valuable information about the retinal surface, its inability to visualize deeper structures limits its diagnostic capabilities [8]. FA, despite offering detailed vascular imaging, requires intravenous contrast agents and can cause adverse reactions in some patients [9]. The introduction of Optical Coherence Tomography (OCT) in the 1990s marked a significant advancement, enabling cross-sectional imaging of retinal layers with micrometer-level resolution [10]. However, conventional OCT’s limitation in visualizing blood flow dynamics created a crucial gap in understanding vascular pathologies.

The advent of Optical Coherence Tomography Angiography (OCTA) has transformed ophthalmological diagnosis by enabling detailed visualization of retinal vasculature without contrast agents, offering superior visualization of the microvasculature compared to traditional fluorescein angiography [11]. The growing adoption of OCTA has led to an exponential increase in imaging data complexity and volume, making manual interpretation increasingly time-consuming and challenging for clinicians. A single OCTA scan can generate multiple en face projections and cross-sectional images, requiring careful examination of various vascular layers [12]. While traditional image processing techniques offered initial automated solutions through handcrafted features like vessel density measurements and foveal avascular zone analysis [13], they often fell short in capturing subtle disease patterns and showed limited generalizability across different pathologies [14]. Deep learning, particularly Convolutional Neural Networks (CNNs), has emerged as a powerful tool for automated OCTA analysis, demonstrating remarkable accuracy in disease classification tasks. Recent studies have achieved classification accuracies exceeding 90% for specific retinal conditions using deep learning approaches [15], [16].

Despite these advances, current deep learning approaches face significant limitations in real-world clinical applications. Most successful implementations have been confined to single-institution settings, where data is centrally collected and processed. This constraint stems from several critical challenges: the scarcity of public datasets (with OCTA-500 being one of few available resources) [17], strict medical data-sharing regulations like HIPAA and GDPR, and significant variations in image acquisition parameters across different OCTA devices and imaging protocols [10]. Furthermore, the prevalence of retinal diseases varies significantly across populations and geographic regions [18], leading to inherent data imbalances. These factors have created isolated data silos, hampering the development of robust, generalizable classification models that could benefit from diverse patient populations.

The imperative to develop robust, generalizable AI models while preserving patient privacy has led to a paradigm shift in medical image analysis. Deep learning research has demonstrated that Federated Learning (FL), a distributed learning framework, can effectively address the challenges of cross-institutional model training while maintaining data privacy [19]. FL represents a transformative approach to collaborative AI development where instead of centralizing sensitive patient data, each institution trains models locally and shares only model parameters, thereby maintaining compliance with privacy regulations while leveraging the collective knowledge of multiple healthcare institutions. This approach has shown remarkable success in various medical imaging applications, including chest X-ray analysis [20], brain tumor segmentation [21], and COVID-19 detection [22]. The utility of distributed FL for ophthalmic diagnosis has been explored recently by where FL demonstrated exceptional capability to train generalizable models across multiple institutions.

The unique characteristics of OCTA imaging make it an ideal candidate for federated learning approaches. OCTA data exhibits significant heterogeneity due to variations in scanning protocols, device manufacturers, and image quality across institutions [24]. Additionally, the complex nature of retinal pathologies and their varying representations across different patient populations [25] necessitate diverse training data that no single institution can provide. FL’s ability to leverage distributed datasets while maintaining patient privacy could potentially address these challenges, enabling the development of more robust and generalizable models for OCTA analysis.

In this work, we demonstrate the first FL based pipeline for OCTA based multi-disease diagnosis. Our investigation utilizes a diverse dataset comprising 500 images from the public OCTA-500 dataset [15] and an additional 446 images from a private collection at the University of Illinois Chicago (UIC). The combined dataset covers seven distinct retinal conditions: Age-related Macular Degeneration (AMD), Choroidal Neovascularization (CNV), Central Serous Chorioretinopathy (CSC), Diabetic Retinopathy (DR), Retinal vein occlusion (RVO), Normal, and Other retinal conditions.

We chose these aggregation strategies based on their potential to address the unique challenges of OCTA classification, such as data heterogeneity, class imbalance, and the need for stable and robust models. FedAvg serves as a baseline, while FedAdagrad and FedYogi introduce adaptive learning rates to handle the varying contributions of different clients. FedProx incorporates proximal terms to improve model stability, and FedMRI utilizes a contrastive loss function to enhance feature representation learning.

Our experimental setup involves training a ResNet50 model, pre-trained on ImageNet, using each of the five aggregation strategies. We evaluate the performance of our FL strategies using various metrics, including accuracy, precision, recall, F1 score, and ROC AUC, both at the overall and per-class levels. Additionally, we analyze the convergence behavior of each strategy to gain insights into their learning dynamics.

The main contributions of this work are as follows:

1. A comprehensive evaluation of five federated learning aggregation strategies (FedAvg, FedAdagrad, FedYogi, FedProx, and FedMRI) for OCTA-based retinal disease classification, establishing a framework for privacy-preserving multi-institutional collaboration.
2. An in-depth analysis of the performance of each strategy on a diverse OCTA dataset, comprising both public (OCTA-500) and private (UIC) data, providing insights into their strengths and limitations for multi-class retinal disease classification.
3. A complete pipeline for federated OCTA analysis that addresses real-world challenges, including data heterogeneity, class imbalance, and privacy requirements, serving as a blueprint for future research and clinical applications.

## II. DATASET

### A. Data Source and Description

In this study we have leveraged optical coherence tomography angiography (OCTA) imaging data from two clinical repositories: the publicly available OCTA500 dataset and a private dataset of OCT images provided by the University of Illinois at Chicago (UIC). OCTA provides high-resolution, depth-resolved visualization of retinal and choroidal vasculature which which allows for non-invasive assessment of vascular networks in various retinal layers without the need for contrast agents. This advanced imaging modality enables detailed examination of microvascular abnormalities associated with various ocular diseases, including diabetic retinopathy, age-related macular degeneration, and glaucoma, providing valuable biomarkers for early disease detection and progression monitoring.

The OCTA500 dataset represents a comprehensive collection of optical coherence tomography angiography (OCTA) images designed for research in retinal vascular analysis. Within this dataset, three distinct projection types are available: OCTA(Full), which captures vascular information throughout the entire retinal depth; OCTA ILM-OPL, which isolates vessels from the inner limiting membrane to the outer plexiform layer, primarily highlighting the superficial and deep capillary plexuses; and OCTA OPL-BM, which focuses on vessels between the outer plexiform layer and Bruch’s membrane, emphasizing the choriocapillaris and choroidal vasculature. For our analysis, we exclusively utilized the OCTA(Full) projection maps (also known as En Face Angiography) because they provide the most comprehensive visualization of retinal vasculature without segmentation artifacts that can occur when focusing on specific layers. This holistic approach was essential for maintaining consistency with the UIC dataset, which required additional processing.

The UIC dataset primarily contained conventional OCT scans rather than native angiographic data. To generate comparable OCTA visualizations from these OCT images, we implemented the advanced computational methodologies detailed in [26]. By applying this rigorous transformation to the UIC dataset, we ensured that all vascular representations across both datasets maintained functional equivalence, despite originating from different imaging modalities, thereby establishing a robust foundation for our comparative analyses.

Thus, our combined dataset from the two sources comprised of 456 OCTA images representing seven distinct retinal pathologies in Scenario A, including Age-related Macular Degeneration (AMD), Choroidal Neovascularization (CNV), Central Serous Chorioretinopathy (CSC), Diabetic Retinopathy (DR), Retinal Vein Occlusion (RVO), normal cases and other retinal conditions. The disease distribution reflects real-world prevalence patterns, with DR and normal cases representing the largest subgroups (31.1% and 25.2% respectively), followed by other retinal conditions (21.1%) and AMD (9.4%). Less common pathologies include CSC (3.1%), CNV (2.4%), and RVO (1.8%).

To facilitate comprehensive analysis, we organized our experiments into three distinct scenarios:

- **Scenario A:** All seven classes (AMD, CNV, CSC, DR, Normal, Others, RVO) were maintained separately.
- **Scenario B:** Four classes were used, with CNV cases incorporated into the AMD class, and both CSC and RVO cases excluded.
- **Scenario C:** Four classes were used (AMD, DR, Normal, Others), completely excluding CNV, CSC, and RVO cases.

This structured approach to dataset organization allows for robust model evaluation across varying levels of class granularity while addressing the inherent challenge of class imbalance in medical imaging datasets. The natural class imbalance in our dataset presents both challenges and opportunities for developing robust classification models.

### B. Data Preprocessing

Our preprocessing pipeline ensured standardization while preserving clinically relevant features. The workflow began with intensity normalization to scale pixel values to a standardized range that optimizes neural network training. We then resized all images to 224×224 pixels through a combination of padding and center cropping, ensuring consistent dimensions across the dataset while maintaining central pathological features.

For the training dataset, we implemented a comprehensive augmentation strategy to enhance model generalizability. This included random 90° rotations, controlled zoom variations (±10%), addition of minor Gaussian noise, contrast adjustments, Gaussian smoothing, and histogram shifts. Each augmentation was applied with carefully calibrated probability to maintain clinical relevance while creating sufficient variability for robust learning. Validation and test datasets underwent the same normalization and standardization procedures without augmentations to ensure faithful evaluation of model performance on clinical-quality images.

### C. Dataset Splitting

We employed a statistically rigorous approach to dataset partitioning to enable comprehensive evaluation across three distinct experimental scenarios. For all scenarios, we utilized a two-stage stratified splitting strategy to maintain pathological representation and source distribution across subsets. The first stage allocated 80% of the data to the training set, with the remaining 20% divided equally between validation and test sets (10% each). Crucially, stratification was performed on both disease class and data source (OCTA500 vs. UIC) simultaneously to ensure balanced representation across all splits.

For Scenario A, we combined images from both OCTA500 and UIC datasets, maintaining all original disease labels. Scenario B involved a targeted reclassification approach where CNV cases were relabeled as AMD, which reflects clinical practice given that CNV is a manifestation of exudative AMD rather than a distinct entity. CSC and RVO samples were excluded to focus on more prevalent conditions. Scenario C further refined the disease spectrum by excluding CNV samples entirely, concentrating on a more limited set of well-differentiated pathologies.

In Scenario A (shown in Table I), all seven disease categories were preserved, resulting in 364 images for training (79.8%), and 46 images each (10.1%) for validation and testing. This comprehensive approach allowed us to evaluate model performance across the full spectrum of retinal pathologies, despite the class imbalance challenges presented by less common conditions such as CNV (2.4%), CSC (3.1%), and RVO (1.8%).

**TABLE I:** Distribution OF IMAGES ACROSS DISEASE CATEGORIES IN THE DATASET SPLITS FOR Scenario A.

| Disease | Training Set |  |  | Validation Set |  |  | Test Set |  |  |
| --- | --- | --- | --- | --- | --- | --- | --- | --- | --- |
|  | OCTA | UIC | Total | OCTA | UIC | Total | OCTA | UIC | Total |
| AMD | 34 | 0 | 34 | 4 | 0 | 4 | 5 | 0 | 5 |
| CNV | 9 | 0 | 9 | 1 | 0 | 1 | 1 | 0 | 1 |
| CSC | 11 | 0 | 11 | 2 | 0 | 2 | 1 | 0 | 1 |
| DR | 28 | 95 | 123 | 3 | 12 | 15 | 4 | 12 | 16 |
| Normal | 73 | 29 | 102 | 9 | 4 | 13 | 9 | 4 | 13 |
| Others | 77 | 0 | 77 | 10 | 0 | 10 | 9 | 0 | 9 |
| RVO | 8 | 0 | 8 | 1 | 0 | 1 | 1 | 0 | 1 |
| Total | 240 | 124 | 364 | 30 | 16 | 46 | 30 | 16 | 46 |

In Scenario B (Table II), we implemented a modified four-class structure where CNV cases were incorporated into the AMD category, while CSC and RVO cases were excluded. This resulted in a more balanced class distribution with 345 total images: 43 AMD (including original CNV cases), 123 DR, 102 normal, and 77 in the “others” category. The training, validation, and test sets contained 345, 43, and 43 images, respectively.

**TABLE II:**
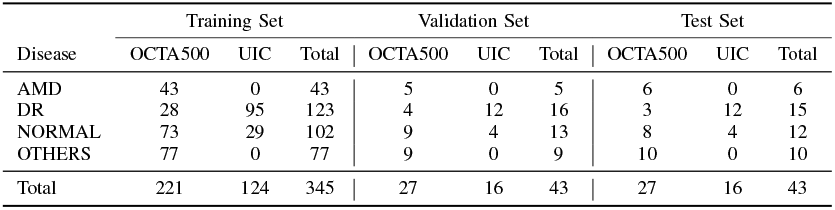
Distribution OF IMAGES ACROSS DISEASE CATEGORIES IN THE DATASET SPLITS FOR Scenario B.

| Disease | Training Set |  |  | Validation Set |  |  | Test Set |  |  |
| --- | --- | --- | --- | --- | --- | --- | --- | --- | --- |
|  | OCTA500 | UIC | Total | OCTA500 | UIC | Total | OCTA500 | UIC | Total |
| AMD | 43 | 0 | 43 | 5 | 0 | 5 | 6 | 0 | 6 |
| DR | 28 | 95 | 123 | 4 | 12 | 16 | 3 | 12 | 15 |
| NORMAL | 73 | 29 | 102 | 9 | 4 | 13 | 8 | 4 | 12 |
| OTHERS | 77 | 0 | 77 | 9 | 0 | 9 | 10 | 0 | 10 |
| Total | 221 | 124 | 345 | 27 | 16 | 43 | 27 | 16 | 43 |

Scenario C (Table III) employed a strict four-class taxonomy (AMD, DR, normal, and others) by completely excluding CNV, CSC, and RVO cases. This produced a streamlined dataset that focused on the most prevalent retinal conditions while maintaining class separation between AMD and CNV. Similar to Scenario B, this configuration utilized 345 total images, with 34 AMD cases representing 9.9% of the dataset.

**TABLE III:**
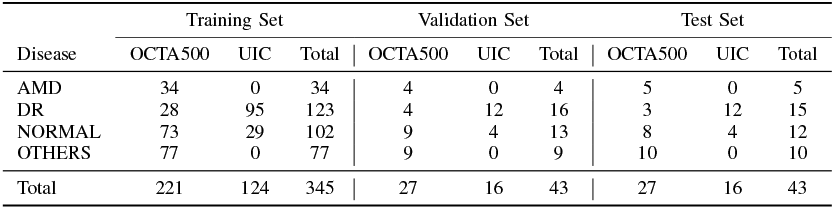
Distribution OF IMAGES ACROSS DISEASE CATEGORIES IN THE DATASET SPLITS FOR Scenario C.

The multi-source composition of our dataset merits particular attention, as it directly influences model learning dynamics. As shown in Fig. 2, DR cases derived from both OCTA500 and UIC sources constitute the largest group across all scenarios, followed by normal cases also obtained from both repositories. The remaining categories -AMD, CNV, CSC, other conditions, and RVO -come exclusively from the OCTA500 dataset. This multi-source approach for common conditions (DR and normal cases) enhances model generalizability, while maintaining comprehensive coverage of rarer pathologies.

**Fig. 1.**
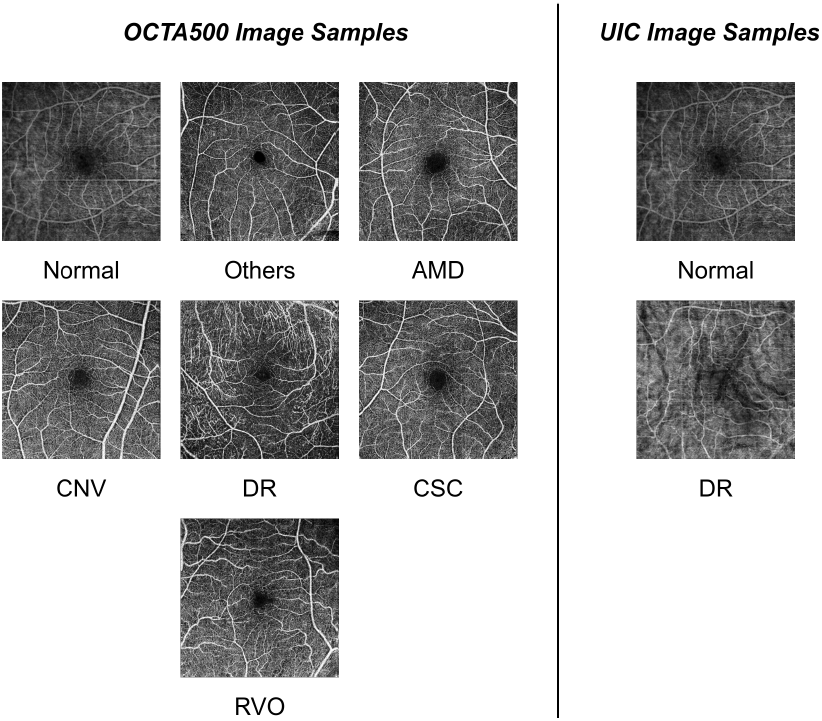
Image samples from each class of both OCTA500 and UIC dataset

**Fig. 2.**
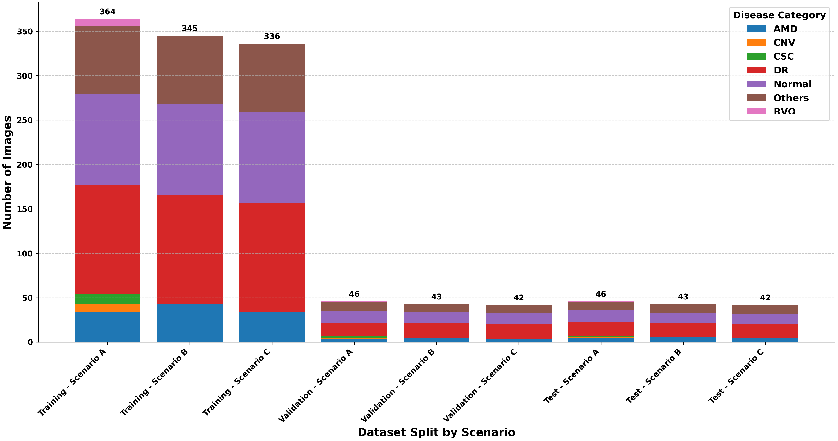
Data distribution bar chart for different scenearios

Our dataset curated for the three experimental scenarios offers several distinctive advantages. The inclusion of seven pathological categories in Scenario A represents broader coverage than most comparable studies, which typically focus on three to four conditions. Furthermore, the multi-center nature of our data, combining images from different institutions and imaging systems, inherently incorporates real-world variability that is crucial for developing robust clinical applications.

## III. METHODOLOGY

### A. Federated Learning Framework

#### 1) Client-Server Model

Our federated learning system implements a client-server architecture as shown in Fig. 3. *K* healthcare institutions collaboratively train a disease classification model while preserving patient data privacy. The central server coordinates the learning process, while distributed clients {*C*_1_, *C*_2_, …, *C*_*K*_} (representing individual healthcare providers) participate with their local OCTA image datasets 𝒟_*k*_.

**Fig. 3.**
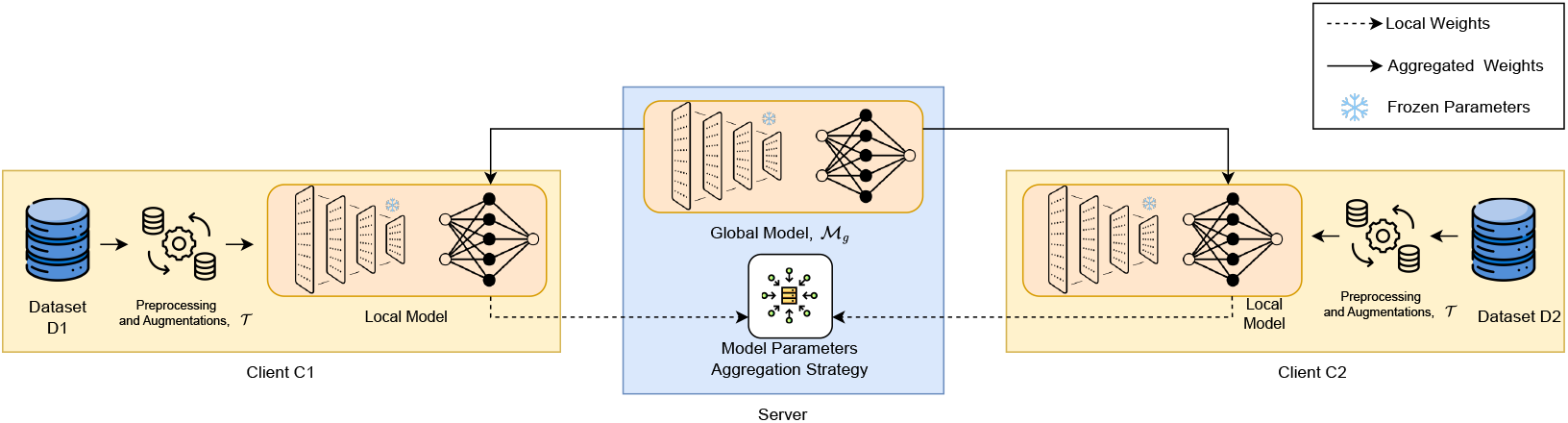
Client-Server architecture of federated learning framework

The system initializes with the server distributing a global model ℳ_*g*_ with parameters *θ*_*g*_—a modified version of ResNet50. We strategically froze most of the model’s convolutional layers, allowing only the final classification layer with parameters *θ*_*f*_ ⊂ *θ*_*g*_ to be trained. This approach leverages pre-existing knowledge about general image features while reducing the computational complexity from 𝒪 (|*θ*_*g*_|) to 𝒪 (|*θ*_*f*_|), where |*θ*_*g*_ |≫|*θ*_*f*_|.

During each communication round *t*, each participating client *C*_*k*_ trains the model using only their local patient data 𝒟_*k*_. To ensure equitable representation, we implemented stratified sampling that maintains a balanced disease distribution across all participating institutions. Formally, for each class *c* ∈ 𝒞;, we ensure *P* (*c* |𝒟_*k*_) ≈ *P* (*c* |𝒟_*j*_) for any clients *k* and *j*. Clients enhance local training with specialized medical image augmentation techniques 𝒯, creating variations through controlled transformations:

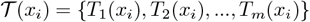

where *x*_*i*_ represents an original image and *T*_*j*_ are transformations including rotations, zooming, and contrast adjustments.

The privacy-preserving aspect of our approach ensures clients never share actual patient images—only the locally trained model parameters 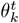 are transmitted back to the server at each round *t*. The server then aggregates these parameters using five federated averaging strategies, like FedAvg, FedProx, FedMRI, FedAdagrad, and FedYogi, each offering different advantages for handling the unique challenges of medical data.

Throughout the training process, we employ a cosine annealing learning rate schedule:

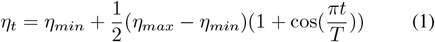

where *η*_*t*_ is the learning rate at round *t, η*_*min*_ and *η*_*max*_ are the minimum and maximum learning rates, and *T* is the total number of rounds.

This collaborative approach enables multiple institutions to contribute to a more robust, generalizable multi-disease classification model characterized by performance metrics like accuracy, f1-score and ROC-AUC score without compromising patient privacy or violating data-sharing regulations.

#### 2) Communication Protocol

Our federated learning system establishes a secure and efficient communication framework between the central server and participating healthcare institutions. The protocol operates through three sequential phases in each federated round: model distribution, decentralized training, and secure aggregation.

During the model distribution phase, the server broadcasts the current global model parameters 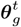 to the selected subset of participating clients {*C*_*k*_|*k* ∈ *S*_*t*_}, where *S*_*t*_ represents the client selection for round *t*. The server accompanies these parameters with configuration metadata that includes the adaptive learning rate *η*_*t*_ derived from our cosine annealing schedule obtained from (1).

This approach ensures that training progressively transitions from exploration to refinement as rounds advance. The server also transmits round-specific metadata, enabling clients to track training progress across the total number of rounds T, such that config_*t*_ = {*η*_*t*_, *t, T*}.

In the decentralized training phase, each participating institution *C*_*k*_ performs local model optimization on their private OCTA image dataset 𝒟_*k*_. Clients execute a fixed number of local epochs (*E* = 3) using the Adam optimizer with the server-specified learning rate *η*_*t*_. The frozen convolutional layers significantly reduce computational complexity while maintaining feature extraction capabilities. The training process can be formally expressed as:

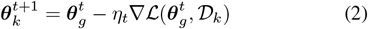

where ℒ represents the cross-entropy loss function evaluated on the local dataset. Upon completion, each client evaluates the updated model on their local validation set to compute performance metrics.

The secure aggregation phase represents the privacy-preserving cornerstone of our approach. Each client *C*_*k*_ transmits only the updated model parameters 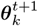 to the server—never revealing actual patient data. The server then performs weighted aggregation based on the relative dataset sizes to update the global model parameters based on the selected federated aggregation strategy.

This weighted approach ensures that institutions with larger and potentially more diverse patient populations contribute proportionally to the global model, while still preserving the influence of smaller specialized centers. After aggregation, the server evaluates the new global model on a centralized validation set to track convergence and performance across various disease classes.

Our protocol design minimizes communication overhead by transmitting only essential model parameters rather than raw medical images, making it practical even with bandwidth constraints. By ensuring patient data never leaves its originating institution, we maintain compliance with privacy regulations while still enabling collaborative improvement of diagnostic capabilities across healthcare systems.

#### 3) Federated simulation environment

To rigorously evaluate our federated learning approach before deployment in actual healthcare settings, we developed a comprehensive simulation environment that replicates real-world conditions while providing experimental control. This environment is built on the Flower framework, which has NVIDIA Flare as a backend. Flower offers the infrastructure needed to simulate multiple distributed clients and their interactions with a central server.

Our simulation accurately models two separate institutional nodes, each with their own distinct patient population. We carefully designed these simulated institutions to reflect real-world medical data challenges—particularly the uneven distribution of rare disease cases. Through stratified sampling, we ensured that each simulated institution has representation across all disease categories as per the experimental scenarios, mirroring the class imbalance typically encountered in clinical practice.

The data is pre-processed which includes standardized sizing, intensity normalization, and domain-specific augmentations that reflect the variations seen in clinical OCTA imaging. This attention to realistic data handling ensures that our simulation results will translate effectively to real-world scenarios.

The environment implements a comprehensive evaluation strategy using a centralized test dataset accessible only to the server. This approach provides an unbiased assessment of how well the collaboratively trained model generalizes to new, unseen patient data. To address the challenges of imbalanced disease prevalence, we calculate macro-averaged metrics that give equal importance to each disease category regardless of its frequency. This is particularly important for ensuring the model performs well on rare but clinically significant conditions.

By balancing realistic medical data challenges with controlled experimental conditions, our simulation environment provides reliable insights that can guide the successful implementation of privacy-preserving federated learning in clinical ophthalmology settings.

### B. Model Architecture

#### 1) Base Network Selection

In this study, we employed ResNet50 [27] as the foundation for our federated learning model. ResNet50 is a variant of the Residual Network architecture that comprises 50 layers, making it sufficiently deep to capture complex features while remaining computationally efficient for distributed training paradigms.

The selection of ResNet50 was motivated by several factors critical to our federated learning implementation. First, the architecture has demonstrated excellent performance across various medical imaging tasks, providing a robust foundation for transfer learning in our specialized OCTA classification scenario. Second, its moderate depth strikes an optimal balance between representational capacity and computational over-head, an essential consideration in federated learning where client-side computational resources may be heterogeneous and constrained [28].

Additionally, ResNet50 offers a well-structured hierarchical feature representation, where initial layers capture low-level features such as edges and textures, while deeper layers progressively extract higher-level semantic features relevant to pathological patterns in OCTA images [29]. This hierarchical structure aligns well with our federated learning strategy, allowing us to freeze earlier layers while fine-tuning the later domain-specific layers, thereby reducing the parameter communication burden across the network while preserving feature extraction capabilities.

#### 2) Feature Extraction Strategy

In our federated learning approach for OCTA image classification, we strategically leveraged the rich feature extraction capabilities of ResNet50 while adapting the network architecture to reduce computational overhead and mitigate overfitting.

A critical aspect of our approach was the decision to freeze all convolutional layers of the pre-trained ResNet50 model. In our implementation, we systematically disabled gradient updates for all parameters in the convolutional backbone, effectively freezing these layers during the training process.

This design choice was motivated by several considerations. First, the lower convolutional layers of networks pre-trained on natural images have been shown to learn general-purpose features that transfer well to medical imaging tasks [30]. Second, freezing these layers substantially reduces the number of trainable parameters, which is particularly advantageous in the federated learning context where parameter transmission between clients and server constitutes a significant communication overhead [31].

By maintaining fixed weights in the feature extraction layers, we ensured consistent feature representations across all participating nodes in the federated network, which promotes stability in the distributed learning process. This approach allowed us to benefit from the rich feature extraction capabilities of ResNet50 while focusing the learning process on adapting only the classification layer to our specific OCTA disease classification task.

#### 3) Classification Layer Design

While maintaining the pre-trained feature extraction layers in a frozen state, we specifically modified the final classification layer of the ResNet50 architecture to accommodate our multi-disease classification task involving OCTA images. We replaced the original ImageNet classifier with a custom classification head by preserving the input dimension of 2048 features from the global average pooling layer but reducing the output dimension to match our 7-class disease classification task for Scenario A and 4-class disease classification task for Scenario B. Crucially, we ensured that only this final fully connected layer remained trainable, while keeping all other network parameters frozen.

Our design deliberately maintains simplicity in the classification head, consisting of only a single linear layer without additional hidden layers. This design choice was motivated by several considerations specific to the federated learning context. First, a simpler classification head with fewer parameters reduces the communication overhead during the model aggregation process. Second, a leaner classifier is less prone to overfitting, which is particularly important given the heterogeneous and potentially limited data available at individual nodes in the federated network.

During training, we employed cross-entropy loss directly on the logits, providing an effective objective function for our multi-class classification task.

#### 4) Model Parameters

The parameter distribution in our adapted ResNet50 architecture plays a crucial role in the efficiency and effectiveness of our federated learning approach for OCTA image classification. By strategically partitioning trainable and non-trainable parameters, we optimized both the learning process and communication overhead.

In our implementation, we froze all parameters in the convolutional layers, which constitute approximately 23.5 million parameters (about 92% of the total network parameters). The only trainable parameters in our model were contained in the final fully connected classification layer, which we resized from the original 2048×1000 configuration to 2048×7 for our seven-class disease classification. This restructured classification layer contains 14,343 parameters, representing only about 0.056% of the total parameters in the network.

This dramatic reduction in trainable parameters offers several advantages in the federated learning context:

1. **Reduced Communication Overhead:** Only the 14,343 trainable parameters need to be exchanged between the federated clients and the server during each round, significantly reducing bandwidth requirements compared to transmitting the full 25.6 million parameters. This is crucial for federated learning systems where communication costs often represent a bottleneck
2. **Improved Convergence:** With fewer trainable parameters, the model requires less data to converge, which is beneficial in medical imaging contexts where data at each node may be limited.
3. **Consistent Feature Extraction:** The frozen feature extraction layers ensure that all clients utilize consistent feature representations, which helps mitigate potential issues arising from data heterogeneity across different nodes.
4. **Efficient Resource Utilization:** The reduced number of trainable parameters decreases the computational requirements for model training at each client, making the approach more feasible for deployment across a range of hardware configurations.

#### 5) Training and Optimization

During training, we employed the Adam optimizer with a cosine annealing learning rate schedule defined by (1) to adaptively adjust parameter updates, providing efficient convergence characteristics suited to federated learning.

The federated learning context introduces an additional dimension to the training process, as the model parameters flow between the server and clients. In each communication round, clients perform local forward and backward passes using their private data, while only the gradients and updated parameters of the trainable classification layer are communicated back to the server. This approach preserves data privacy while enabling collaborative learning across distributed OCTA datasets.

### C. Aggregation Strategies

Federated learning aggregation strategies are critical for addressing the unique challenges inherent in distributed medical image analysis. For our OCTA multi-disease classification task, effective aggregation must balance model performance, data heterogeneity, and the nuanced requirements of ophthalmic diagnosis. In this study, we have implemented and evaluated five different baseline strategies to demonstrate how different aggregation strategies affect the federated model. A brief description about each of them is as follows:

#### Federated Averaging (FedAvg)

FedAvg [32] represents the foundational approach to federated learning. The global model update is computed as a weighted average of local model updates:

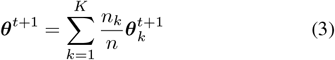

where ***θ***^*t*+1^ represents the global model parameters, *n*_*k*_ is the local dataset size, and *n* is the total dataset size. In our OCTA classification context, FedAvg provides a baseline method that efficiently aggregates knowledge across clinical sites while preserving patient privacy. However, its performance may be affected by the inherent data heterogeneity across different ophthalmic imaging centers, as variations in imaging protocols and patient demographics can create non-IID data distributions.

#### Federated Proximal (FedProx)

To address the challenges of model divergence common in medical imaging datasets, FedProx introduces a proximal term that constrains local model updates:

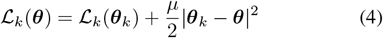

where *µ* is a proximal regularization parameter. This approach is particularly valuable for our OCTA dataset, where each clinical site may have different disease prevalence patterns and imaging characteristics. By preventing significant deviation from the global model, FedProx improves stability and convergence when confronted with the heterogeneous nature of retinal disease manifestations across different patient populations.

#### Federated Learning for MR Image reconstruction (FedMRI)

For our complex medical image classification task, we also implemented FedMRI, which introduces contrastive learning principles to federated aggregation:

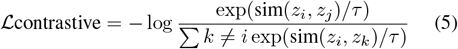

where sim(*z*_*i*_, *z*_*j*_) measures the similarity between model representations. This strategy is particularly promising for OCTA imaging, as it helps the model learn consistent feature representations across different client datasets, potentially improving the recognition of subtle disease biomarkers that may manifest differently across varied imaging conditions and patient populations.

#### Federated Adagrad

Medical image classification often requires nuanced parameter updates across different model components. FedAdagrad extends adaptive gradient methods to federated learning with per-parameter learning rate adaptation:

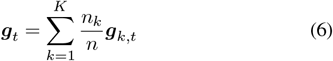

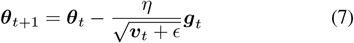

where ***g***_*t*_ represents the aggregated gradient, and ***v***_*t*_ accumulates squared gradients. For our ResNet50-based model, this adaptive learning approach is potentially beneficial for fine-tuning feature extraction from OCTA images, where different convolutional layers may require varying update magnitudes to effectively capture disease-specific patterns across different retinal layers and structures.

#### Federated Yogi

To further enhance the adaptive capabilities of our federated model, we implemented FedYogi, which improves upon Adam-like algorithms with a more sophisticated gradient accumulation mechanism:

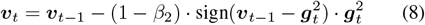

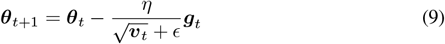

This approach is particularly relevant for our seven-class OCTA disease classification task, where some pathologies (like diabetic retinopathy) may be more prevalent than others. FedYogi’s noise-reduction properties help prevent over-fitting to dominant classes while maintaining responsiveness to gradient updates for rarer conditions, potentially improving the overall diagnostic accuracy across the disease spectrum.

### D. Implementation Details

#### 1) Experimental Setup

Our federated learning framework was implemented using Flower [33] (version 1.13.1), an open-source federated learning framework that provides robust infrastructure for federated learning experimentation. All models were developed using PyTorch 2.2.1 and TorchVision 0.17.1, with custom implementations of several federated optimization strategies including FedAvg [32], FedProx [34], FedMRI [35], FedAdagrad [36], and FedYogi [36].

#### 2) Federated Learning Configuration

We implemented our federated learning experiments using a simulation-based approach to emulate a practical multi-center clinical scenario. The federated learning environment was configured with 2 simulated clients, each representing a distinct medical center with its own local dataset. For each round of federated training, all available clients were selected to participate, ensuring maximum utilization of available data while maintaining privacy constraints.

The server was configured to run for 50 federated rounds, with a minimum requirement of 2 available clients for each round to proceed. We implemented and compared multiple federated optimization strategies. After each global aggregation, the server evaluated the updated global model on separate validation datasets from each client, as well as on a centralized test dataset. This approach allowed us to monitor both client-specific and overall model performance throughout the training process.

To ensure fair comparison between different federation strategies, we maintained identical client selection, data partitioning, and communication protocols across all experiments, varying only the optimization and aggregation algorithms. This controlled experimental design enabled us to isolate the impact of each federated optimization strategy on model performance, convergence rate, and generalization capabilities across heterogeneous medical imaging data.

#### 3) Evaluation Metrics

To comprehensively assess the performance of our federated learning models for multi-disease classification in OCTA images, we implemented a diverse set of evaluation metrics that address the challenges of class imbalance often present in medical imaging datasets. Our primary metric was macro-averaged accuracy, which calculates the average of per-class accuracies, giving equal weight to each disease class regardless of its prevalence in the dataset. This approach was particularly important for our multi-disease classification task, as it prevented the model evaluation from being dominated by more common conditions. In addition to macro-averaged accuracy, we also calculated the F1-score using macro-averaging to maintain fairness across classes. For each disease class,also we tracked individual metrics including per-class accuracy, F1-score, and class-specific ROC-AUC values. This granular analysis allowed us to identify specific disease classes that might benefit from additional optimization or data augmentation strategies.

For model comparison and selection, we utilized the area under the receiver operating characteristic curve (ROC-AUC) with a one-vs-rest approach for multi-class classification. The ROC-AUC metric provided a threshold-independent assessment of our models’ discriminative capabilities across all disease classes. To handle potential numerical instabilities, particularly with classes having limited samples, we incorporated a small epsilon value (1e-7) to prevent division by zero and ensure robust metric calculation.

We evaluated model performance at three levels: on each client’s validation dataset to assess local performance, on the combined validation data to measure generalization across clients, and on a separate centralized test dataset to evaluate performance on unseen data. This multi-level evaluation approach allowed us to monitor both the federated learning process and the final model’s clinical applicability. For each experiment, we saved per-round metrics and automatically identified the best-performing model configuration based on centralized test accuracy, facilitating systematic comparison between different federated optimization strategies.

## IV. RESULTS

In this section, we present a comprehensive evaluation of our federated learning approaches for multi-class disease classification using OCTA images. We conducted experiments across three distinct scenarios using data from two difference sources-publicly available OCTA500 dataset and private dataset from UIC. The three scenarios are as follows:

- Scenario A with all 7 classes (AMD, DR, Normal, Others, CNV, CSC, RVO)
- Scenario B with 4 modified classes (CNV considered as AMD, and CSC/RVO dropped)
- Scenario C with 4 classes (CNV, CSC, and RVO completely dropped)

For each scenario, we compare centralized training (using all available data), standalone client models (trained only on individual client datasets), and five baseline federated learning approaches (FedAvg, FedProx, FedMRI, FedYogi, and FedAdagrad). This experimental design allows us to analyze the trade-offs between data privacy preservation and diagnostic accuracy, a critical consideration in contemporary medical AI research.

### A. Global Test Set Performance

#### 1) Scenario A: Seven-Class Classification

Table IV presents the accuracy results for all methods in the seven-class classification scenario. The centralized model achieved 65.22% accuracy. The standalone models for Client1 and Client2 achieved 60.87% and 58.70% accuracy, respectively. Among the federated learning approaches, FedMRI demonstrated the best performance with 60.87% accuracy, matching the performance of Client1’s standalone model.

**TABLE IV:** Accuracy (%) COMPARISON ACROSS SCENARIOS ON Global Test Set (Note: Scenario A INCLUDES ALL 7 CLASSES (AMD, DR, Normal, Others, CNV, CSC, RVO); Scenario B HAS 4 CLASSES WITH CNV CONSIDERED AS AMD AND CSC/RVO DROPPED; Scenario C HAS 4 CLASSES WITH CNV, CSC, AND RVO COMPLETELY DROPPED. The BEST FEDERATED LEARNING PERFORMANCE IN EACH SCENARIO IS HIGHLIGHTED IN BOLD.)

| Method | Scenario A<br>(7 Classes) | Scenario B<br>(4 Mod. Classes) | Scenario C<br>(4 Classes) |
| --- | --- | --- | --- |
| Centralized | 65.22 | 68.18 | 69.77 |
| Client1 Standalone | 60.87 | 68.18 | 67.44 |
| Client2 Standalone | 58.70 | 61.36 | 69.77 |
| FedAvg | 46.70 | 65.91 | **72.09** |
| Fedprox | 30.25 | 68.18 | **72.09** |
| FedMRI | **60.87** | 61.36 | **72.09** |
| FedYogi | 28.26 | 45.45 | 37.21 |
| FedAdagrad | 45.65 | 29.55 | 30.23 |

In contrast, FedProx (30.25%), FedYogi (28.26%), and FedAdagrad (45.65%) showed lower performance compared to other approaches.

When evaluating with ROC-AUC (Table V), FedAvg achieved the highest performance among federated methods at 79.12%, close to the centralized model’s performance (79.55%). For F1-score (Table VI), FedAvg substantially outperformed all other methods, including the centralized approach, with a score of 46.08% compared to 32.22% for the centralized model.

**TABLE V:** ROC-AUC (%) COMPARISON ACROSS SCENARIOS ON Global Test Set.

| Method | Scenario A<br>(7 Classes) | Scenario B<br>(4 Mod. Classes) | Scenario C<br>(4 Classes) |
| --- | --- | --- | --- |
| Centralized | 79.55 | 84.50 | 86.21 |
| Client1 Standalone | 67.44 | 80.33 | 73.10 |
| Client2 Standalone | 75.00 | 79.64 | 86.88 |
| FedAvg | **79.12** | 81.36 | 86.12 |
| Fedprox | 76.52 | 80.68 | 86.46 |
| FedMRI | 70.70 | **81.64** | **87.17** |
| FedYogi | 72.75 | 69.15 | 72.66 |
| FedAdagrad | 71.74 | 61.90 | 69.50 |

**TABLE VI:** F1-Score (%) COMPARISON ACROSS SCENARIOS ON Global Test Set.

| Method | Scenario A<br>(7 Classes) | Scenario B<br>(4 Mod. Classes) | Scenario C<br>(4 Classes) |
| --- | --- | --- | --- |
| Centralized | 32.22 | 60.64 | 53.48 |
| Client1 Standalone | 28.33 | 63.85 | 52.92 |
| Client2 Standalone | 26.10 | 45.86 | 54.42 |
| FedAvg | **46.08** | 57.84 | **62.41** |
| Fedprox | 27.60 | 55.51 | 56.05 |
| FedMRI | 27.48 | **53.54** | 56.17 |
| FedYogi | 6.30 | 33.97 | 20.83 |
| FedAdagrad | 16.00 | 11.40 | 11.61 |

#### 2) Scenario B: Four Modified Classes

In Scenario B, where CNV cases were considered as AMD and CSC/RVO samples were dropped resulting in 4 classes, we observed a general improvement in performance across most methods (Table IV). The centralized model achieved 68.18% accuracy, while Client1 and Client2 standalone models achieved 68.18% and 61.36%, respectively. FedProx matched the centralized model’s performance at 68.18%.

For ROC-AUC (Table V), FedMRI achieved the highest score among federated methods at 81.64%, outperforming both FedAvg (81.36%) and FedProx (80.68%). Similarly for F1-score (Table VI), FedMRI achieved the best performance among federated methods at 53.54%, though it did not surpass the standalone Client1 model (63.85%).

#### 3) Scenario C: Four Classes with Dropped Samples

Scenario C, where CNV, CSC, and RVO samples were completely dropped resulting in 4 classes, showed the highest overall performance across all scenarios (Table IV). The centralized model achieved 69.77% accuracy, while Client1 and Client2 standalone models achieved 67.44% and 69.77%, respectively. All three federated methods—FedAvg, FedProx, and FedMRI—achieved 72.09% accuracy, outperforming both the centralized and standalone approaches.

For ROC-AUC (Table V), FedMRI again achieved the highest score at 87.17%, surpassing the centralized model (86.21%) and Client2 standalone model (86.88%). For F1-score (Table VI), FedAvg achieved the highest performance at 62.41%, substantially outperforming both centralized (53.48%) and standalone approaches (52.92% and 54.42%).

### B. Client-Specific Performance Analysis

The client-specific validation results in Table VII show varying performance patterns across clients. For Client1, FedMRI performed well across all scenarios (50.00%, 57.14%, and 71.43%). In contrast, Client2 showed different responses to federated methods depending on the scenario—with FedMRI performing best in Scenario A (45.83%), and FedAvg in Scenarios B and C (77.27% and 66.67%). In Scenario B, FedAvg improved Client2’s performance by 13.63% compared to their standalone model (77.27% vs. 63.64%).

**TABLE VII:** Accuracy (%) ON EACH Client’s Validation Sets ACROSS SCENARIOS.

| Method | Scenario A |  | Scenario B |  | Scenario C |  |
| --- | --- | --- | --- | --- | --- | --- |
|  | Client1 | Client2 | Client1 | Client2 | Client1 | Client2 |
| Client1 Standalone | 66.67 | 45.83 | 52.38 | 77.27 | 57.14 | 52.38 |
| Client2 Standalone | 58.33 | 37.50 | 57.14 | 63.64 | 52.38 | 57.14 |
| FedAvg | 28.16 | 30.60 | 57.14 | **77.27** | 71.43 | 66.67 |
| Fedprox | 31.02 | 23.57 | 47.62 | 68.18 | **76.19** | 61.90 |
| FedMRI | **50.00** | **45.83** | **57.14** | 68.18 | 71.43 | 52.38 |
| FedYogi | 29.17 | 25.00 | 52.38 | 40.91 | 33.33 | 28.57 |
| FedAdagrad | 25.00 | 37.50 | 28.57 | 31.82 | 28.57 | 28.57 |

In Scenario B, FedMRI and FedAvg tied for the best performance on Client1’s validation set (57.14%), while FedAvg outperformed other methods on Client2’s validation set (77.27%).

For Scenario C, Fedprox achieved the highest accuracy on Client1’s validation set (76.19%), while FedAvg performed best on Client2’s validation set (66.67%).

## V. DISCUSSION

Our comprehensive evaluation reveals several key findings that advance the field of federated learning for medical imaging. First, federated learning can match or even outperform centralized training in certain scenarios, particularly when the class structure is simplified (Scenarios B and C). This result challenges the prevailing assumption that federated learning necessarily entails a performance penalty compared to centralized approaches. Second, we observed that FedMRI was specifically designed to handle the unique characteristics of medical imaging data by incorporating domain-specific regularization terms that account for the high dimensionality and spatial correlations in OCTA images, explaining its strong ROC-AUC performance. Meanwhile, FedAvg’s aggregation mechanism appears to achieve a more balanced precision-recall trade-off, potentially because it inherently mitigates the impact of class imbalance across institutions. Third, the underperformance of adaptive methods like FedYogi and FedAdagrad can be attributed to challenges in converging to a consensus model when confronted with the high degree of statistical heterogeneity present in multi-institutional retinal imaging data.

The comparative analysis of methods reveals nuanced insights about federated learning in medical imaging contexts. Figure 4 illustrates the comparative performance of centralized, standalone, and various federated learning methods across three data distribution scenarios. While the centralized model achieved the highest accuracy in Scenario A, the performance gap between federated and centralized approaches was relatively small (4.35% for FedMRI), representing a favorable privacy-utility trade-off. Most remarkably, in Scenario C, federated methods outperformed centralized training by 2.32%, contradicting previous studies that positioned federated learning as necessarily sacrificing performance for privacy. FedAvg’s unexpected superiority in F1-score can be attributed to its aggregation mechanism producing a more balanced precision-recall trade-off than other methods, making it less susceptible to overfitting on majority classes that could dominate in centralized training. This better balancing of class representation is particularly valuable in clinical contexts where rare conditions must be identified with similar reliability as common ones. The gradient distributions in retinal imaging likely exhibit high variance across institutions due to differences in imaging protocols, patient demographics, and disease presentation variability, explaining why adaptive methods struggle with the sparse representation of certain disease features in distributed datasets.

**Fig. 4.**
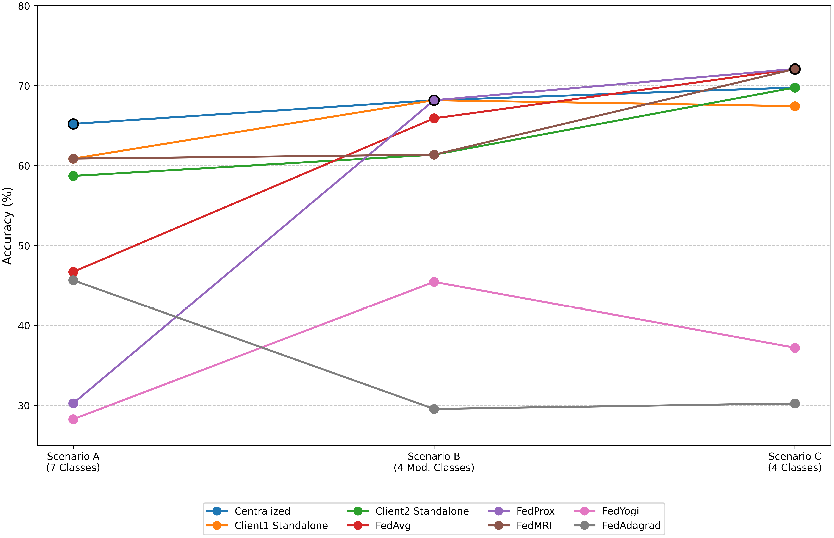
Accuracy comparison across different scenarios on Global Test Set

From a clinical perspective, our results present several actionable insights. First, the strong performance of federated methods in Scenario C demonstrates that healthcare institutions can achieve superior diagnostic performance through federated collaboration without compromising patient data privacy—a critical consideration under regulations like HIPAA and GDPR. This performance gain likely stems from the federated methods’ ability to learn complementary features from diverse institutional data while avoiding overfitting to institution-specific biases that may affect the centralized model. Second, the high ROC-AUC values achieved by FedMRI indicate reliable discrimination between disease classes, which is critical in clinical applications where false negatives can have severe consequences. Third, the variable performance across clients highlights that institutional factors—potentially including patient demographics, disease prevalence, and imaging equipment—significantly impact which federated approach is optimal, suggesting that personalized or adaptive federation strategies might be necessary for multi-center clinical deployments.

Despite these promising results, several limitations must be acknowledged. First, our two-client federation represents a simplified case of institutional collaboration; in real-world implementations, heterogeneity would likely increase with more participating centers, potentially introducing additional challenges for model convergence. Second, while our evaluation across three class configurations provides useful insights, clinical deployment would require validation with prospective cases presenting varying disease severities and comorbidities not fully represented in our current datasets. Third, although our evaluation metrics are comprehensive from a machine learning perspective, they may not fully capture clinically relevant performance characteristics such as sensitivity to early-stage disease or ability to detect disease progression over time. Fourth, computational efficiency considerations—particularly important for resource-constrained health-care settings—warrant dedicated investigation, as the communication overhead of federated methods may present implementation challenges in bandwidth-limited environments. Finally, our results on client validation sets show variable performance across institutions, suggesting that model personalization strategies may be necessary to ensure consistent diagnostic quality across all participating healthcare providers.

In summary, our results demonstrate that federated learning approaches, particularly FedMRI, FedAvg, and FedProx, can effectively leverage multi-institutional OCTA imaging data for disease classification while maintaining data privacy. We found that when the class structure is simplified, the proximal term in FedProx effectively addresses data heterogeneity by constraining local updates to remain close to the global model, preventing client drift—a significant challenge in federated medical imaging. Meanwhile, FedMRI’s specialized architecture incorporates medical imaging-specific inductive biases, allowing it to better capture the subtle discriminative features that distinguish between retinal pathologies. These findings suggest that specialized approaches are needed for medical imaging that differ from those effective for natural image classification, and provide a foundation for future research into personalized federated learning approaches that can adapt to the specific characteristics of each participating institution’s data.

## VI. CONCLUSION

This study provides a comprehensive evaluation of federated learning strategies for multi-disease classification using Optical Coherence Tomography Angiography (OCTA) images across different healthcare institutions. Our investigation across three distinct classification scenarios demonstrates that federated learning can effectively balance the dual imperatives of diagnostic accuracy and patient privacy protection in ophthalmology.

The comparative analysis of five federated aggregation strategies—FedAvg, FedProx, FedMRI, FedAdagrad, and FedYogi—reveals nuanced performance patterns that have significant implications for clinical deployment across all three experimental scenarios. In Scenario A (seven-class classification), FedMRI demonstrated the strongest performance among federated approaches (60.87% accuracy), approaching the centralized model’s performance (65.22%) while FedAvg achieved the highest F1-score (46.08%), substantially outperforming the centralized approach (32.22%). In Scenario B (four modified classes with CNV incorporated into AMD), FedProx matched the centralized model’s accuracy (68.18%), while FedMRI achieved the highest ROC-AUC (81.64%). Most notably, in our simplified four-class scenario (Scenario C), FedAvg, FedProx, and FedMRI all achieved 72.09% accuracy, surpassing the centralized training approach (69.77%). These findings challenge the conventional wisdom that federated learning necessarily entails a performance sacrifice compared to centralized approaches, instead suggesting that collaborative learning can enhance model robustness by incorporating diverse institutional perspectives while maintaining data sovereignty.

Our results also highlight the differential strengths of various federated methods across evaluation metrics and scenarios. FedMRI consistently demonstrated superior ROC-AUC performance (reaching 87.17% in Scenario C and 81.64% in Scenario B), indicating its strong discriminative capability across disease classes—a critical factor in clinical settings where missed diagnoses can have serious consequences. FedAvg showed strong performance in ROC-AUC for Scenario A (79.12%) while exhibiting remarkable strength in F1-score metrics (46.08% in Scenario A and 62.41% in Scenario C), suggesting its effectiveness in balancing precision and recall across classes, particularly in the presence of class imbalance. The consistent underperformance of adaptive methods like FedYogi and FedAdagrad across all scenarios indicates that medical imaging requires specialized federated approaches that differ from those effective in natural image classification.

The client-specific performance analysis reveals that institutional factors significantly influence which federated approach yields optimal results, with Client1 consistently benefiting from FedMRI while Client2 showed variable preferences across scenarios. This observation underscores the importance of considering institutional data characteristics when deploying federated learning systems in multi-center clinical settings.

From a clinical perspective, our work demonstrates that healthcare institutions can leverage federated learning to enhance diagnostic capabilities without compromising patient privacy—addressing a critical barrier to AI adoption in medical imaging. The strong performance in specific disease classification scenarios suggests that targeted implementations focusing on clinically relevant subsets of conditions may yield the best results in practice.

Several promising directions for future research emerge from our findings. First, developing adaptive federation strategies that dynamically adjust to institutional data characteristics could further enhance performance across heterogeneous healthcare networks. Second, exploring personalization techniques to optimize client-specific performance while maintaining global model quality represents an important area for investigation. Third, extending our evaluation to larger institutional networks would provide insights into the scalability of these approaches for widespread clinical deployment. Finally, integrating domain-specific knowledge through medical priors or expert-guided feature selection could potentially enhance the interpretability and diagnostic accuracy of federated models.

In conclusion, our comprehensive evaluation demonstrates that federated learning, particularly using FedMRI, FedAvg, and FedProx strategies, offers a viable approach for privacy-preserving collaboration across healthcare institutions for OCTA-based disease classification. By enabling institutions to collectively improve diagnostic models without sharing sensitive patient data, federated learning addresses a fundamental challenge in medical AI deployment. Our findings not only advance the technical understanding of federated learning in medical imaging but also provide practical insights for implementing these approaches in clinical ophthalmology settings, potentially accelerating the translation of AI advances into improved patient care.

## Data Availability

All data produced in the present study are available upon reasonable request to the authors. Moreover, OCTA500 dataset is publicly available online.

https://ieee-dataport.org/open-access/octa-500

## ACKNOWLEDGMENT

The preferred spelling of the word “acknowledgment” in American English is without an “e” after the “g.” Use the singular heading even if you have many acknowledgments. Avoid expressions such as “One of us (S.B.A.) would like to thank … .” Instead, write “F. A. Author thanks … .” In most cases, sponsor and financial support acknowledgments are placed in the unnumbered footnote on the first page, not here.

---

**First A. Author** (Fellow, IEEE) and all authors may include biographies. Biographies are often not included in conference-related papers. This author is an IEEE Fellow. The first paragraph may contain a place and/or date of birth (list place, then date). Next, the author’s educational background is listed. The degrees should be listed with type of degree in what field, which institution, city, state, and country, and year the degree was earned. The author’s major field of study should be lower-cased.

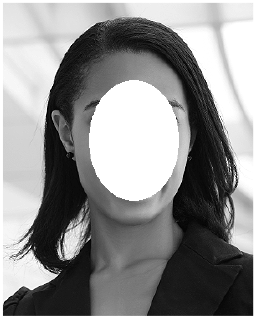

The second paragraph uses the pronoun of the person (he or she) and not the author’s last name. It lists military and work experience, including summer and fellowship jobs. Job titles are capitalized. The current job must have a location; previous positions may be listed without one. Information concerning previous publications may be included. Try not to list more than three books or published articles. The format for listing publishers of a book within the biography is: title of book (publisher name, year) similar to a reference. Current and previous research interests end the paragraph.

The third paragraph begins with the author’s title and last name (e.g., Dr. Smith, Prof. Jones, Mr. Kajor, Ms. Hunter). List any memberships in professional societies other than the IEEE. Finally, list any awards and work for IEEE committees and publications. If a photograph is provided, it should be of good quality, and professional-looking.

**Second B. Author**, photograph and biography not available at the time of publication.

**Third C. Author Jr**. (Member, IEEE), photograph and biography not available at the time of publication.

